# Day by day symptoms following positive and negative PCR tests for SARS-CoV-2 in non-hospitalised health-care workers: a 90-day follow-up study

**DOI:** 10.1101/2021.03.02.21252437

**Authors:** Kent J. Nielsen, Jesper Medom Vestergaard, Vivi Schlünssen, Jens Peter Bonde, Kathrine Agergård Kaspersen, Karin Biering, Ole Carstensen, Thomas Greve, Karoline Kærgaard Hansen, Annett Dalbøge, Esben Meulengracht Flachs, Sanne Jespersen, Mette Lausten Hansen, Susan Mikkelsen, Marianne Kragh Thomsen, Jacob Dvinge Redder, Else Toft Würtz, Lars Østergaard, Christian Erikstrup, Henrik Albert Kolstad

## Abstract

**Background:** Little is known about the long-term course of symptoms for mild coronavirus disease 2019 (COVID-19) when accounting for symptoms due to other causes. We aimed to compare symptoms day by day for non-hospitalised individuals who tested positive and negative with polymerase chain reaction for severe acute respiratory syndrome coronavirus 2 (SARS-CoV-2).

**Methods:** We followed 210 test-positive and 630 individually matched test-negative health-care workers of the Central Denmark Region up to 90 days after the test, April-June 2020. They daily reported seven COVID-19 related symptoms. Symptom courses were compared graphically and by conditional multivariable logistic regression.

**Results:** Thirty % of test-positive and close to zero of test-negative participants reported a reduced sense of taste and smell during all 90 days of follow-up (adjusted odds ratio [aOR] 86.07, 95% CI 22.86-323). Dyspnoea was reported by an initial 20% of test-positive with a gradual decline to about 5% after 30 days without ever reaching the level of the test-negative participants (aOR 6.88, 95% CI 2.41-19.63). Cough, headache, sore throat, muscle aches, and fever were temporarily more prevalent among the test positive participants, but after 30 days, no increases were seen. Women and participants aged 45 years or older tended to be more susceptible to SARS-CoV-2 infection.

**Conclusion:** Prevalence of long-lasting reduced sense of taste and smell is highly increased after being diagnosed with mild COVID-19. This pattern is also seen for dyspnoea at a low level but not for cough, sore throat, headache, muscle ache or pain, or fever.

**Key messages:**

- Reduced sense of taste and smell is present at a highly increased level of 30% during 90 days after testing positive for severe acute respiratory syndrome coronavirus 2 (SARS-Cov-2).
- Test-positive participants experience dyspnoea persistently more often than test-negative participants but affect only few.
- The prevalence of cough, sore throat, headache, muscle ache or pain, and fever following a positive test reach the level seen after a negative test within 30 days.
- Women and participants aged 45 years or older tend to be more susceptible to symptoms following SARS-CoV-2 infection.

## Introduction

Severe acute respiratory syndrome coronavirus 2 (SARS-CoV-2) has affected most countries during the last year leading to the coronavirus disease 2019 (COVID-19) pandemic.^1^ The clinical manifestations of acute SARS-CoV-2 infection range from asymptomatic, over mild symptoms to life-threatening infection with compromised respiratory capacity and organ failure. Most patients hospitalised with COVID-19 present fatigue, fever, cough, dyspnoea, musculoskeletal pain, headache, and reduced sense of taste and smell.^2,3^ A high proportion still have symptoms, particularly fatigue, anosmia, sleep difficulties, and musculoskeletal pain after recovery.^4-6^ There is increasing concern about the long-term consequences and a post-COVID-19 syndrome is being discussed.^7-9^

Uncontrolled data from the general population and non-hospitalised COVID-19 patients with mild disease indicate that a high proportion suffer from SARS-CoV-2 related symptoms several weeks after diagnosis.^10-15^ Prospective follow-up studies of non-hospitalised COVID-19 patients including a reference group accounting for symptoms not attributable to SARS-CoV-2 are warranted.^16^ The few studies comparing symptom courses of test positive with test negative non-hospitalised participants show increased occurrence of reduced sense of taste and smell and several other symptoms that persist for several weeks and months after a positive SARS-CoV-2 test.^17,18^ We aimed to compare day by day symptoms of SARS-CoV-2 PCR test-positive and test-negative non-hospitalised health-care workers up to 90 days after the test.

## Methods

### Design and study setting

We carried out a prospective follow up study of health care workers and other occupational groups from all hospitals in the Central Denmark Region from April 24 until June 30, 2020.

### Participants

All hospital employees were invited by e-mail to report COVID-19 related symptoms day by day. Participants tested by PCR for SARS-CoV-2 from March 11 until June 30, 2020, at any of the regional hospitals or public test centres, were identified in the Central Denmark Region business intelligence system. We included those with at least one daily report on symptoms from the day being tested and onwards. We excluded those hospitalised for COVID-19 for more than 24 hours because our focus was non-hospitalised individuals.

### PCR-test for SARS-CoV-2 RNA

National surveillance in Denmark of SARS-CoV-2 infection assessed by reverse transcription PCR-based detection of viral RNA in nasopharyngeal and oropharyngeal swabs was initiated on March 2, 2020.^19,20^ Until March 11, only symptomatic individuals returning from high-risk areas and symptomatic contacts could be tested. From March 12, also individuals with severe symptoms, individuals at-risk because of high age or comorbidity, or with critical functions could be tested. From April 1, further individuals with mild symptoms; and from April 21, close contacts regardless of symptoms had the opportunity to be tested. From May 18, all adults have been offered testing. Since April 21, all patients have been tested before being admitted to the hospital or undergoing high-risk procedures during outpatient visits. PCR analysis for SARS-CoV-2 RNA was performed at the Clinical Microbiology Department at Aarhus University Hospital with detection of the ORF-1a/b and E-gene (commercial assay) or exclusively the E-gene, and at the national test-facilities at the TestCentre Denmark, Statens Serum Institut, with detection of the E-gene, both in-house PCRs in accordance with the Charité protocol recommended by the WHO.^21,22^ Automated RNA extraction was performed at both facilities. Internal negative and positive controls were included in both the RNA extraction step and in the reverse transcription PCR step.

### Questionnaire

After giving informed consent, participants received a short baseline questionnaire and then a short text message on their mobile phone or by e-mail every day at 3:30 pm linking to a questionnaire regarding the presence (yes, no) of the following symptoms within the previous 24 hours: cough, sore throat, headache, fever, muscle aches and pains, dyspnoea, and reduced or lost sense of taste and smell (available in supplementary data). Participants could respond within 24 hours from receiving the message and could resume reporting if skipping one or more days. Smoking status was collected in the baseline questionnaire.

### Other data

Information on occupation, sex, and age was provided by the business intelligence institution of the Central Denmark Region.

### Statistical analyses

We followed participants from the date of the first completed questionnaire after the first positive test, else from the first negative test until the date of the last questionnaire, 90 days after being tested, or June 30, whichever came first. No participants had a positive test after a negative test during the follow-up period.

Because the indication for being tested, testing rate, and infection rate in the study population changed rapidly over time (Supplementary Figure S1) we for each participant tested positive, randomly selected three referents with replacement among participants tested negative matching on sex and testing date (+/- 2 days). The three-fold number of referents was defined by the maximum allowed within the narrowest strata. When selecting referents, we avoided crossing the specific dates where indications for being tested changed as specified above.

For test-positive and test-negative participants, we computed the prevalence of the seven symptoms as well as any of the symptoms for each day of follow-up. We plotted the prevalences and smoothed the curves with local three-degree polynomial kernels. Standard error based 95% confidence intervals (95% CI) were obtained based on 100 bootstrap samples, resampling among the test-positive participants and repeating the matching of test-negative participants and the smoothing procedure.

We estimated odds ratios (OR) of any symptom and the seven specific symptoms by test result (positive, negative) for three time periods (0-30, 31-60, 61-90 days) since the test by conditional logistic regression matched by sex and testing date as specified above. We assessed if sex modified the symptom prevalence among test-positive relative to test-negative participants by including an interaction term between test result and sex (man, woman). We also assessed the possible modifying effect of age (<45, ≥45 years, the median age) and testing date (≤April 7, >April 7, the median testing date) similarly. We assessed selection bias, i.e. if test-positive and test-negative participants’ responding on the questionnaire on a given day were modified by the presence of symptoms the previous day, in a model that included test result, any symptom (present, absent), the interaction term between the two, and responding on the questionnaire (yes, no). The conditional logistic regression models were adjusted for age (<30, 30-39, 40-49, 50-59, and ≥60 years), except analyses of effect modification by age, occupation (nursing staff, medical doctors, biomedical laboratory scientists, medical secretaries, and other), smoking (current, previous, and never), unless else specified. Overall odds ratios for the entire follow up period were furthermore adjusted by time since test (0-30, 31-60, 61-90 days). The covariates were decided on a priori. Confidence intervals were obtained by bootstrapping as described above. Data handling and statistical analyses were performed in Stata 16.1.

## Results

Between April 23 and May 5, 32 413 health-care workers and administrative personnel were invited to participate in the day by day symptom reporting, and 12 115 (37·4%) accepted. Between March 11 and June 30, 215 respondents were tested PCR-positive for SARS-CoV-2, and 3421 were tested PCR-negative. Five of the test-positive and four of the test-negative participants were hospitalised for >24 hours on the suspicion of COVID-19 and were excluded. Among the remaining 3417 test-negative participants, we randomly selected 630 referents matched on sex and testing date and representing 447 individuals. The study population then included 210 test-positive and 630 test-negative participants. Two referents were selected five times, the maximum number of repeats observed. Data from a mean of 50 test-positive and 164 test-negative participants were included for day 0-30, 128 and 431 for day 31-60, and 87 and 300 for day 61-90 since the test.

The sex and testing date distributions were identical for test-positive and test-negative participants, as expected due to the matched design, and only minor differences in age, smoking habits, and time from test to responding on the first questionnaire were seen (table 1). The mean daily response rate declined from 80.9% and 79.1% during day 0-30 for the test-positive and test-negative participants to 54.6% and 63.1% during day 61-90, respectively. The nursing staff was relatively more prevalent compared to other occupations (administrative, service, and technical staff, social workers, and other less prevalent occupations) with limited patient contact among the test-positive participants. During the first days after being tested, about 80% of the test-positive and 75% of the test-negative participants reported at least one of the seven symptoms (figure 1). Ninety days later, these prevalences had gradually declined to about 40% and 10%, respectively. This corresponded with four-fold increased odds ratios for the complete follow-up period (adjusted odds ratio [aOR] 3.79, 95% CI 2.54-5.66) and for each of the three periods since testing (table 2).

**Table 1.** Characteristics of 210 SARS-Cov-2 test-positive and 630 SARS-Cov-2 test-negative participants matched on sex and testing date

|  |  |  |
| --- | --- | --- |
|  | **Positive test** | **Negative test** |
| **Sex** |  |  |
| Women | 177 (84.3%) | 531 (84.3%) |
| Men | 33 (15.7%) | 99 (15.7%) |
| **Testing date** |  |  |
| March 12-31 | 57 (27.1%) | 171 (27.1%) |
| April 1-20 | 135 (64.3%) | 405 (64.3%) |
| April 21 - May 17 | 18 (8.6%) | 54 (8.6%) |
| May 18 - June 30 | 0 | 0 |
| **Age, years** |  |  |
| <30 | 33 (15.7%) | 58 (9.2%) |
| 30-39 | 49 (23.3%) | 153 (24.3%) |
| 40-49 | 64 (30.5%) | 221 (35.1%) |
| 50-59 | 49 (23.3%) | 146 (23.2%) |
| ≥60 | 15 (7.1%) | 52 (8.3%) |
| **Number of days from the test to first questionnaire response** |  |  |
| 0-30 | 173 (82.4%) | 526 (83.5%) |
| 31-60 | 37 (17.6%) | 103 (16.3%) |
| 61-90 | 0 (0.0%) | 1 (0.2%) |
| **Mean daily response rate since the test (range)** |  |  |
| Day 0-30 | 80.9% (69-100) | 79.1% (70-94) |
| Day 31-60 | 63.9% (57-73) | 71.1% (64-78) |
| Day 61-90 | 54.6% (47-60) | 63.1% (58-70) |
| **Occupation** |  |  |
| Nursing staff | 140 (66.7%) | 290 (46.0%) |
| Medical doctors | 38 (18.1%) | 111 (17.6%) |
| Biomedical laboratory scientists | 8 (3.8%) | 37 (5.9%) |
| Medical secretaries | 5 (2.4%) | 39 (6.2%) |
| Other<sup>a</sup> | 19 (9.0%) | 153 (24.3%) |
| **Smoking** |  |  |
| Current smoker | 10 (4.8%) | 29 (4.6%) |
| Previous smoker | 60 (28.6%) | 204 (32.4%) |
| Never smoker | 140 (66.7%) | 397 (63.0%) |
| <sup>a</sup>Administrative, service and technical staff, social workers, and other less prevalent occupations |  |  |

**Table 2.** Adjusted odds ratios of seven symptoms by SARS-CoV-2 test result and time since the test

|  | Time since the test |  |  |  |  |  |  |  |  |  |  |  |  |  |  |  |
| --- | --- | --- | --- | --- | --- | --- | --- | --- | --- | --- | --- | --- | --- | --- | --- | --- |
|  | Day 0-30 |  |  |  |  | Day 31-60 |  |  |  |  | Day 61-90 |  |  |  |  | Day 0-90 |
|  | Positive test (173 participants) 1552 daily recordings<sup>a</sup> |  | Negative test (526 participants) 5096 daily recordings<sup>a</sup> |  | Adjusted odds ratio (95% CI)<sup>b</sup> | Positive test (181 participants) 3828 daily recordings<sup>a</sup> |  | Negative test (581 participants) 12 920 daily recordings<sup>a</sup> |  | Adjusted odds ratio (95% CI)<sup>b</sup> | Positive test (148 participants) 2608 daily recordings<sup>a</sup> |  | Negative test (515 participants) 8997 daily recordings<sup>a</sup> |  | Adjusted odds ratio (95% CI)<sup>b</sup> | Adjusted odds ratio (95% CI)<sup>b</sup> |
|  | n | % | n | % |  | n | % | n | % |  | n | % | n | % |  |  |
| Any symptom | 862 | 55.5 | 1426 | 28.0 | 4.18<br>(2.63-6.62) | 1689 | 44.1 | 2604 | 20.2 | 3.59<br>(2.37-5.44) | 1003 | 38.5 | 1319 | 14.7 | 4.59<br>(2.44-8.64) | 3.79<br>(2.54-5.66) |
| Reduced or lost sense of taste and smell | 491 | 31.6 | 92 | 1.8 | 57.16<br>(16.71-195) | 1120 | 29.3 | 217 | 1.7 | 62.66<br>(15.15-259) | 745 | 28.6 | 77 | 0.9 | 226.38<br>(160.23-319) | 86.07<br>(22.86-323) |
| Dyspnoea | 119 | 7.7 | 81 | 1.6 | 10.93<br>(2.29-52.10) | 179 | 4.7 | 133 | 1.0 | 6.76<br>(1.79-25.47) | 92 | 3.5 | 48 | 0.5 | 6.27<br>(0.53-73.45) | 6.88 (2.41-19.63) |
| Headache | 227 | 14.6 | 531 | 10.4 | 1.53<br>(1.00-2.33) | 337 | 8.8 | 907 | 7.9 | 1.34<br>(0.84-2.13) | 172 | 6.6 | 480 | 5.3 | 1.21<br>(0.59-2.49) | 1.32<br>(0.81-2.18) |
| Cough | 340 | 21.9 | 641 | 12.6 | 2.19<br>(1.10-4.37) | 405 | 10.6 | 1023 | 7.9 | 1.27<br>(0.75-2.15) | 106 | 4.1 | 492 | 5.5 | 0.81 (0.32-2.08) | 1.33<br>(0.81-2.18) |
| Sore throat | 149 | 9.6 | 439 | 8.6 | 1.33<br>(0.77-2.34) | 115 | 3.0 | 661 | 5.1 | 0.60<br>(0.28-1.27) | 72 | 2.8 | 364 | 4.0 | 0.61<br>(0.21-1.77) | 0.82<br>(0.46-1.48) |
| Muscle ache or pain | 78 | 5.0 | 180 | 3.5 | 1.96<br>(0.74-5.18) | 129 | 3.4 | 314 | 2.4 | 1.40<br>(0.56-3.49) | 94 | 3.6 | 205 | 2.3 | 2.57<br>(0.65-10.14) | 1.69<br>(0.79-3.59) |
| Fever | 14 | 0.9 | 23 | 0.5 | 3.26<br>(0.81-13.10) | 5 | 0.1 | 8 | 0.1 | 1.88<br>(0.42-8.40) | 0 | 0 | 13 | 0.1 | .. | 2.78<br>(0.93-8.34) |

<sup>a</sup>n represents number of responses stating the presence of the specified symptom within the last 24 hours and % represents the proportion of all responses.
<sup>b</sup>Adjusted odds ratios with 95% confidence intervals (CI) are obtained from conditional logistic regression models with 1:3 matching of test-positive with test-negative participants on testing date (+/- 2 days) and sex (man, woman). Models are adjusted for age (<30, 30-39, 40-49, 50-59, and ≥60 years), smoking (current, previous, and never), and occupation (nursing staff, medical doctors, biomedical laboratory scientists, medical secretaries, and other), except for analyses of reduced or lost sense of taste and smell and fever due to unstable estimates that did not provide valid confidence intervals by bootstrapping. Adjusted odds ratios for day 0-90 were furthermore adjusted by time since the test (day 0-30, 31-60, and 61-90). The conditional logistic regression models provide instantaneous odds ratios that cannot be estimated from the period cumulative numbers and percentages of the table.

**Figure 1.**
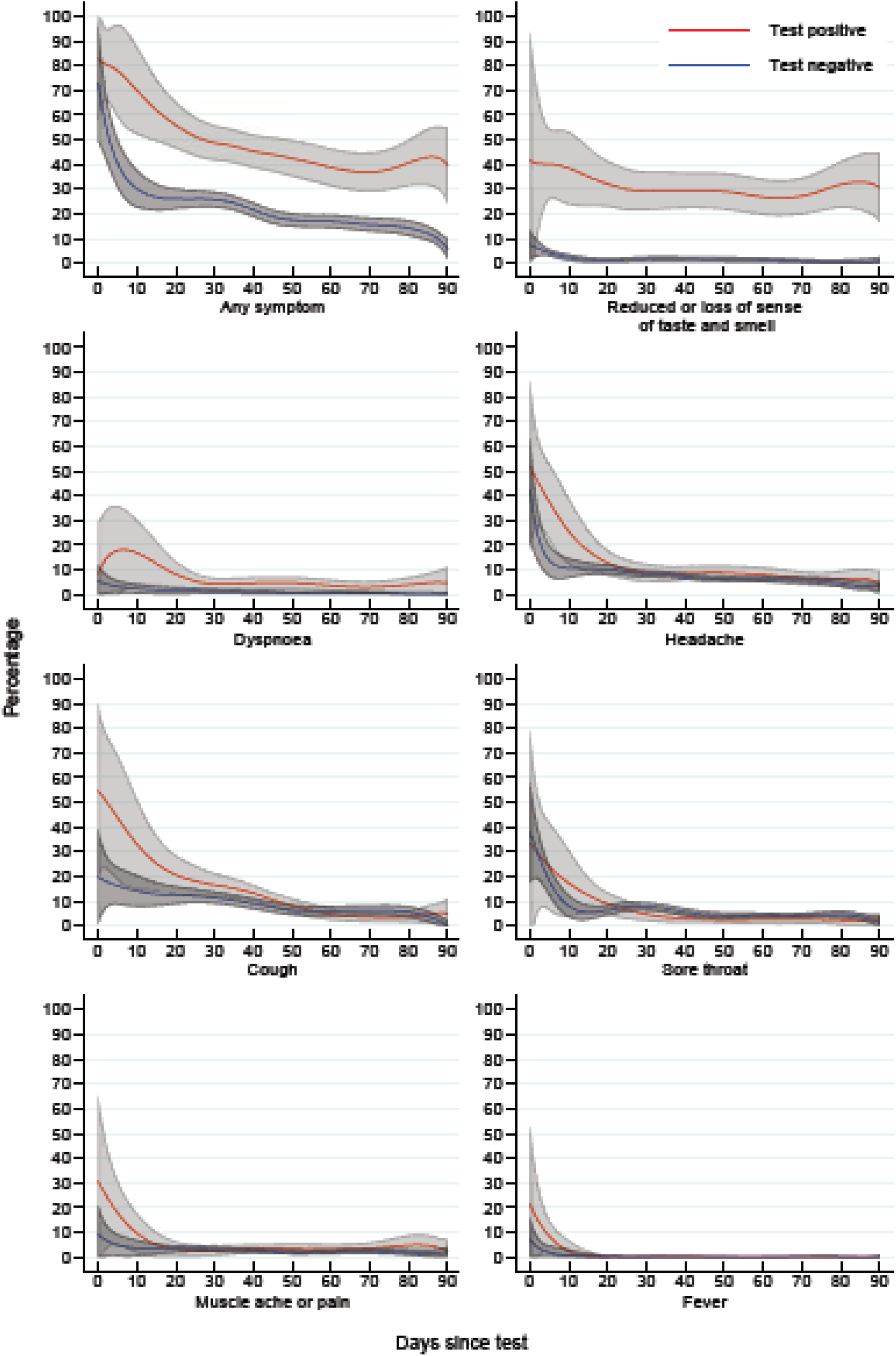
Symptom prevalences (%) by days since SARS-CoV-2 PCR test. 210 participants tested positive and 630 participants tested negative for SARS-CoV-2 and individually matched on sex and testing date. Confidence intervals are depicted by the shadowed areas

Reduced or lost sense of taste and smell was consistently reported by 30% of the test-positive participants, except for a somewhat higher level during the initial days (figure 1). Almost none of the test-negative participants reported these symptoms. The odds ratio tended to increase by time since testing and the overall estimate was 80-fold increased (aOR 86.07, 95% CI 22.86-323, table 2). Dyspnoea was reported by an initial 20% of test-positive participants and declined gradually to about 5% after 30 days without ever reaching the level of the test-negative participants (figure 1). During the first 30 days of follow-up, the odds ratio was 11-fold increased (aOR 10.93, 95% CI 2.29-52.10) compared to test-negative participants. This ratio was reduced during subsequent days and an overall adjusted odds ratio of 6.88 (95% CI 2.41-19.63) was observed.Half of the test-positive and 15% of the test-negative participants reported cough during the initial days (figure 1). The adjusted odds ratio for the first 30 days was 2.19 (95% CI 1.10-4.37). After 30 days, no difference between the two test results was observed. At the time of the test, sore throat, muscle aches or pain, and fever were reported by 35%, 30%, and 20% of the test-positive and this was slightly more than among the test-negative participants. No differences were indicated for these symptoms after 30 days of follow-up.

Test-positive participants aged 45 years or older showed an overall five-fold increased odds ratio (aOR 5.37, 95% CI 2.84-10.14) of any symptom compared with same-age test-negative participants (table 3). The corresponding odds ratio obtained among participants <45 years of age was 2.43, (95% CI 1.42-4.16) and the P-value of the interaction term was 0.07. Similar patterns were seen for day 31-60 and day 61-90, but not for day 0-30. When breaking this analysis down by the seven symptoms, it appeared that this effect modification by age was primarily seen for reduced or lost sense of taste and smell and headache more than 30 days after the test (Supplementary Table S1).

**Table 3.** Adjusted odds ratios of any symptom^a^ by SARS-CoV-2 test result, age, sex, testing date, and time since the test

|  |  |  |  |  |  |  |  |  |  |  |  |  |  |  |  |  |
| --- | --- | --- | --- | --- | --- | --- | --- | --- | --- | --- | --- | --- | --- | --- | --- | --- |
|  | Time since the test |  |  |  |  |  |  |  |  |  |  |  |  |  |  |  |
|  | Day 0-30 |  |  |  |  | Day 31-60 |  |  |  |  | Day 61-90 |  |  |  |  | Day 0-90 |
|  | Positive test and recording of any symptom<sup>b</sup> |  | Negative test and recording of any symptom<sup>b</sup> |  | Adjusted odds ratio (95% CI)<sup>c</sup> | Positive test and recording of any symptom<sup>b</sup> |  | Negative test and recording of any symptom<sup>b</sup> |  | Adjusted odds ratio (95% CI)<sup>c</sup> | Positive test and recording of any symptom<sup>b</sup> |  | Negative test and recording of any symptom<sup>b</sup> |  | Adjusted odds ratio (95% CI)<sup>c</sup> | Adjusted odds ratio (95% CI)<sup>c</sup> |
|  | n | % | n | % |  | n | % | n | % |  | n | % | n | % |  |  |
| Age |  |  |  |  |  |  |  |  |  |  |  |  |  |  |  |  |
| <45 years | 378 | 50.3 | 640 | 28.3 | 4.18 (2.20-7.97) | 558 | 32.6 | 1340 | 22.3 | 2.17 (1.20-3.93) | 301 | 27.8 | 727 | 17.4 | 1.96 (0.89-4.31) | 2.43 (1.42-4.16) |
| ≥45 years | 484 | 60.4 | 786 | 27.8 | 4.33 (2.15-8.72) | 1131 | 53.4 | 1264 | 18.3 | 5.04 (2.63-9.66) | 702 | 46.0 | 592 | 12.3 | 8.50 (3.33-21.67) | 5.37 (2.84-10.14) |
| P value<sup>d</sup> |  |  |  |  | 0.95 |  |  |  |  | 0.08 |  |  |  |  | 0.02 | 0.07 |
| Sex |  |  |  |  |  |  |  |  |  |  |  |  |  |  |  |  |
| Women | 782 | 56.8 | 1293 | 28.3 | 4.26 (2.60-6.98) | 1569 | 47.5 | 2271 | 20.2 | 4.16 (2.73-6.36) | 954 | 42.9 | 1098 | 14.6 | 5.51 (2.92-10.39) | 4.38 (2.90-6.60) |
| Men | 80 | 45.7 | 133 | 25.0 | 3.51 (0.87-14.8) | 120 | 23.0 | 333 | 20.1 | 1.03 (0.31-3.48) | 49 | 12.8 | 221 | 15.1 | 1.10 (0.03-40.0) | 1.44 (0.48-4.36) |
| P value<sup>d</sup> |  |  |  |  | 0.80 |  |  |  |  | 0.03 |  |  |  |  | 0.38 | 0.05 |
| Testing date |  |  |  |  |  |  |  |  |  |  |  |  |  |  |  |  |
| ≤April 7, 2020 | 229 | 63.4 | 482 | 36.9 | 5.34 (2.47-11.54) | 1070 | 48.6 | 1840 | 24.4 | 3.43 (2.14-5.48) | 777 | 41.8 | 980 | 14.9 | 5.49 (2.74-11.00) | 4.11 (2.51-6.74) |
| >April 7, 2020 | 633 | 53.1 | 944 | 24.9 | 3.90 (2.16-7.06) | 619 | 38.1 | 764 | 14.2 | 3.83 (1.83-8.01) | 226 | 30.3 | 339 | 14.0 | 2.79 (0.87-9.00) | 3.59 (1.90-6.77) |
| P value<sup>d</sup> |  |  |  |  | 0.55 |  |  |  |  | 0.79 |  |  |  |  | 0.30 | 0.73 |
| <sup>a</sup>Any symptom includes reduced or lost sense of taste and smell, dyspnoea, cough, headache, sore throat, muscle aches or pain, and fever.<br><sup>b</sup>n represents number of responses stating the presence of any symptom within the last 24 hours and % represents the proportion of all responses.<br><sup>c</sup>Adjusted odds ratios with 95% confidence intervals (CI) are obtained from conditional logistic regression models with 1:3 matching of test-positive with test-negative participants on testing date (+/- 2 days) and sex (man, woman). Models include test result (positive, negative), age (<45 years, ≥45 years), smoking (current, previous, and never), occupation (nursing staff, medical doctors, biomedical laboratory scientists, medical secretaries, and other), and the interaction term between test result and age, test-result and sex, or test result and testing date (≤April 7, >April 7). Adjusted odds ratios for day 0-90 are furthermore adjusted by time since the test (day 0-30, 31-60, and 61-90). The conditional logistic regression models provide instantaneous odds ratios that cannot be estimated from the period cumulative numbers and percentages of the table. Confidence intervals are obtained by bootstrapping.<br><sup>d</sup>The P value is the p value of the interaction term between test result and age, test result and sex, and test result and testing date. |  |  |  |  |  |  |  |  |  |  |  |  |  |  |  |  |

Women who tested positive reported any symptom more often than women who tested negative (aOR 4.38, 95% CI 2.90-6.60) while this was not the case for men (aOR 1.44, 95% CI 0.48-4.36, table 3) and the P-value of the interaction term was 0.05. A similar pattern was seen for day 30-60 and day 61-90 but not for day 0-30. After day 30, much higher prevalences of reduced sense of taste and smell were seen for test-positive relative to test-negative women than for test-positive relative to test-negative men (Supplementary table S2). A similar pattern was suggested for dyspnoea but at a lower level.

Early vs late testing date (≤April 7 vs >April 7) did not modify the association between a positive test and any symptom (table 3).

Among study participants reporting any symptom the previous day, those who tested positive did not respond more often on the present-day questionnaire than those tested negative (aOR 0.93, 95% CI 0.75-1.15, table 4). This was also the case among participants reporting no symptoms the previous day (aOR 1.15, 95% CI 0.88-1.51). The P value of the interaction term was 0.19 and indicated that responding to the questionnaire did not depend on the presence of symptoms the previous day and test result.

**Table 4.**
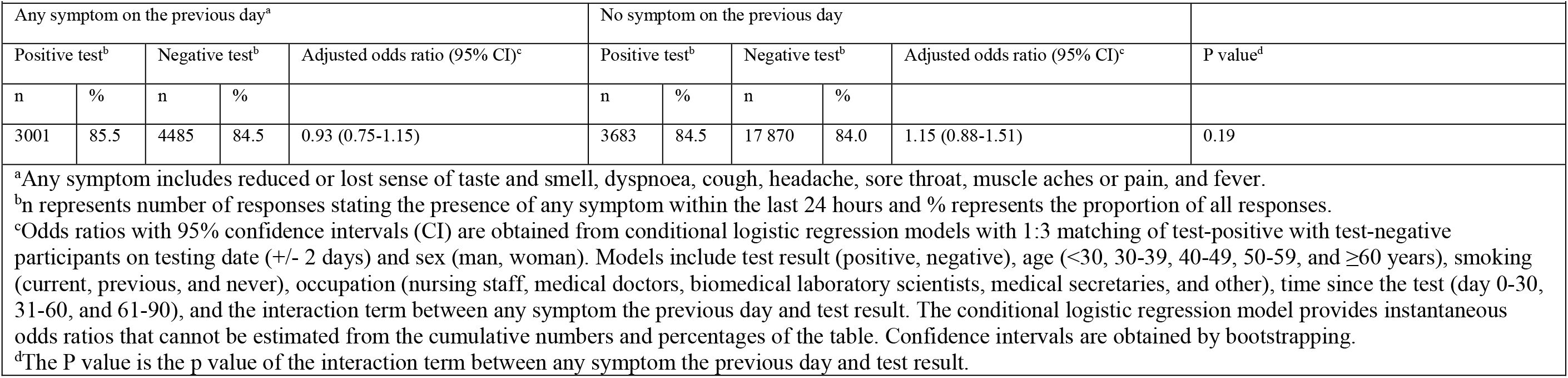
Odds ratios of responding on present-day questionnaire by any symptoms the previous day and SARS-CoV-2 PCR test result

|  |  |  |  |  |  |  |  |  |  |  |
| --- | --- | --- | --- | --- | --- | --- | --- | --- | --- | --- |
| Any symptom on the previous day<sup>a</sup> |  |  |  |  | No symptom on the previous day |  |  |  |  |  |
| Positive test<sup>b</sup> |  | Negative test<sup>b</sup> |  | Adjusted odds ratio (95% CI)<sup>c</sup> | Positive test<sup>b</sup> |  | Negative test<sup>b</sup> |  | Adjusted odds ratio (95% CI)<sup>c</sup> | P value<sup>d</sup> |
| n | % | n | % |  | n | % | n | % |  |  |
| 3001 | 85.5 | 4485 | 84.5 | 0.93 (0.75-1.15) | 3683 | 84.5 | 17 870 | 84.0 | 1.15 (0.88-1.51) | 0.19 |
| <sup>a</sup>Any symptom includes reduced or lost sense of taste and smell, dyspnoea, cough, headache, sore throat, muscle aches or pain, and fever.<br><sup>b</sup>n represents number of responses stating the presence of any symptom within the last 24 hours and % represents the proportion of all responses.<br><sup>c</sup>Odds ratios with 95% confidence intervals (CI) are obtained from conditional logistic regression models with 1:3 matching of test-positive with test-negative participants on testing date (+/- 2 days) and sex (man, woman). Models include test result (positive, negative), age (<30, 30-39, 40-49, 50-59, and ≥60 years), smoking (current, previous, and never), occupation (nursing staff, medical doctors, biomedical laboratory scientists, medical secretaries, and other), time since the test (day 0-30, 31-60, and 61-90), and the interaction term between any symptom the previous day and test result. The conditional logistic regression model provides instantaneous odds ratios that cannot be estimated from the cumulative numbers and percentages of the table. Confidence intervals are obtained by bootstrapping.<br><sup>d</sup>The P value is the p value of the interaction term between any symptom the previous day and test result. |  |  |  |  |  |  |  |  |  |  |

Women, middle-aged employees, and nursing staff were more prevalent in the study population than in the source population (Supplementary table S3).

## Discussion

### Key results

Nearly one-third of SARS-Cov-2 test-positive and close to zero of test-negative participants reported reduced sense of taste and smell during all 90 days of follow-up. Dyspnoea was reported by an initial 20% of test-positive participants and declined gradually to about 5% after 30 days without ever reaching the level of the test-negative participants. Cough, headache, sore throat, muscle aches, and fever were temporarily higher among the test positive participants, but after 30 days, no increases were seen. Women tended to be more susceptible to reduced sense of taste and smell and dyspnoea, and participants aged 45 years or older to reduced sense of taste and smell and headache beyond 30 days.

### Limitations and strengths

The major limitation is the study participants’ awareness of their test results before reporting symptoms, which is expected to have inflated reporting among the test-positive participants. Such an effect is probably strongest for loss of sense of taste and smell that has contracted public awareness worldwide and nationally.^23,24^ Another limitation is only few observations during the first weeks after the test. Hereby the study primarily addresses the course of symptoms after the initial acute phase of the infection.

The prospective design with daily collection of symptom reports that provides information with high temporal resolution is a major strength and makes us able to depict the courses of symptoms day by day. Another strength is the inclusion of a reference group of test-negative participants recruited within the same population as the test-positive health-care workers and tested with the same kit at the same time. This allows us to take symptoms among the test-positive participants not attributed to SARS-CoV-2 infection into consideration and also to account for rapid changes in indications for testing, infection rate, and testing rate in the population. Matching on sex and adjustment for age, smoking, and occupation is expected to have further reduced potential confounding.

Our access to the results of all SARS-CoV-2 tests conducted by the Health Authorities on all samples obtained in the Central Denmark Region during the study period independently of the participants should ensure inclusion of all tested participants and precludes selection or information bias related to testing status. One-third of the invited employees volunteered for symptom reporting and among them, one third was PCR tested. Relatively more nursing staff participated in the study compared to other occupations with limited patient contact. This should have increased the proportion of test-positive participants, but not have affected the validity of symptom comparisons between test-positive and test-negative participants.^25^

Indication for a SARS-CoV-2 PCR-test, testing and infection rates changed during the course of the study, and for that reason, we matched participants individually on testing date. We observed no difference in the association between the test result and any symptom among participants tested early vs late during spring 2020, indicating that matching had fulfilled the purpose. We observed no indications that responding to the questionnaire on a given day depended on test results and symptoms the previous day, and this indicates no differential attrition.

### Comparison with other studies

Our finding of a highly and constantly increased prevalence of reduced or lost sense of taste and smell among the SARS-CoV-2 test-positive compared with the test-negative participants is partly in accordance with two recent reports from general population samples in Israel and the US including few or no participants hospitalised for COVID-19.^18^ Both studies showed initial prevalences among the test-positive participants comparable with ours, but prevalences declined to about 5% after 20 days and to 14% after 90 days, respectively. In both studies, symptom prevalences of lost sense of taste and smell among test-negative participants were constantly close to zero during follow-up in line with our findings. High initial prevalences of altered sense of smell and taste of 60-90% followed by steep recovery rates of 41-87% during 30 days of follow-up have been reported in non-hospitalised patient series.^12,13^ Similar findings were also seen in a follow-up study of mainly COVID-19 outpatients examined with olfactory and gustatory psychophysical tests.^26^ The first days after the test, 85% had taste and smell dysfunction, which gradually declined to 7% 60 days later.

A five-fold increased prevalence of dyspnoea among test-positive compared with test-negative participants (16% vs 3%) 90 days after the test has been reported and is in line with our findings but at a higher absolute level.^18^ Others have reported a constant level of dyspnoea of 30% among test-positive participants during 14-21 days of follow-up in a study that included no reference group, as well as minor difference between test-positive and test-negative participants during 20 days of follow-up.^10,17^

Increased prevalences of cough, sore throat, body aches, and fever among test-positive relative to test-negative individuals 90 days after the test,^18^ high prevalences of the same symptoms among test-positive individuals 14-21 days after the test,^10^ as well as no relative symptom increase in test-positive individuals 20 days after the test, have been reported.^19^ The latter finding being in line with ours. It should be stressed that our study accounted for the testing date, and this may explain some of the inconsistencies between earlier findings and ours.^17,18^

Our data suggest that women and older individuals are more susceptible than men and younger individuals to suffer from COVID-19 related symptoms. There is ample evidence of men being more severely affected by COVID-19 than women, and our contradictory findings may point towards explanations other than SARS-CoV-2 infection per se.^27^

## Conclusion

We observe a highly increased prevalence of long-lasting reduced or lost sense of taste and smell among participants diagnosed with mild COVID-19. This pattern is also seen for dyspnoea at a low level but not for cough, sore throat, headache, muscle ache or pain, or fever. Women and participants aged 45 years or older tend to be more susceptible to SARS-CoV-2 infection.

## Ethics approval

This study was approved by the Danish Data Protection Agency (Jnr 1-16-02-150-20) and the Danish Patient Safety Authority (Jnr. 1-45-70-25-20). The Regional Scientific Ethics Committee of the Central Denmark Region approved that this study did not require scientific ethical approval (Jnr. 1-10-72-1-20).

## Data availability

The data underlying this article will be shared on reasonable request to the corresponding author.

## Data Availability

The data that support the findings of this study are available from the corresponding author upon reasonable request.

## Author contributions

HAK, KJN, JMV, VS, KB, OC, KKH, AD, MLH, ETW, and TG planned the study and collected the data. TG and MKT were responsible for PCR tests. HAK, JMV, KJN, VS, JPB, KAK, and KHK analysed data. HAK and KJN drafted the manuscript. KJN, AD, KB, ETW, KAK, KKH, and HAK did the literature search. JMV designed figures. All authors interpreted and critically revised the manuscript and approved the final version. HAK, KJN, and JMV verified the underlying data.

## Supplementary data to

**Figure S1.**
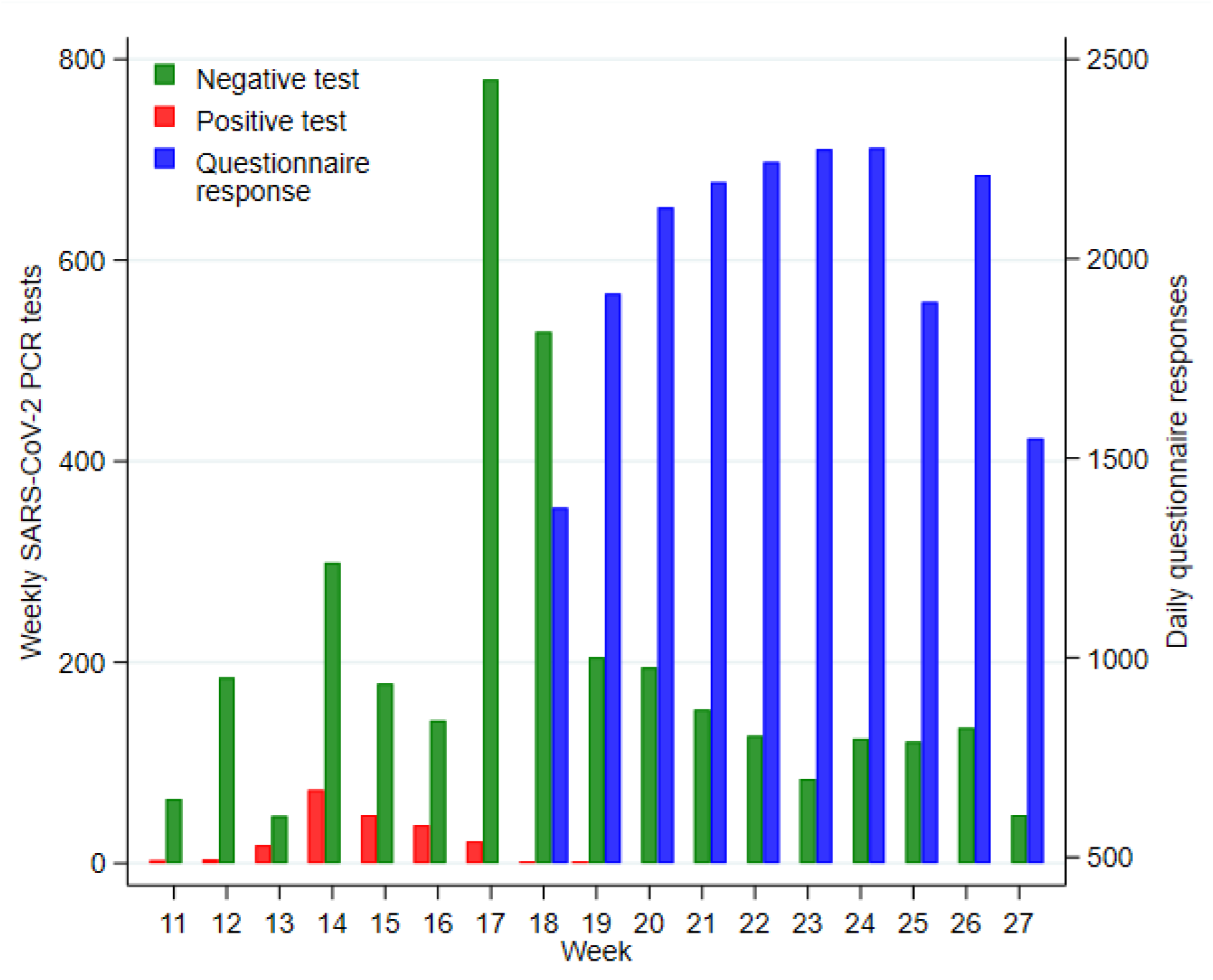
Weekly SARS-CoV-2 PCR tests and daily questionnaire responses by week 11-27, 2020, for 210 SARS-CoV-2 test-positive and 3417 test-negative participants

**Table S1:**
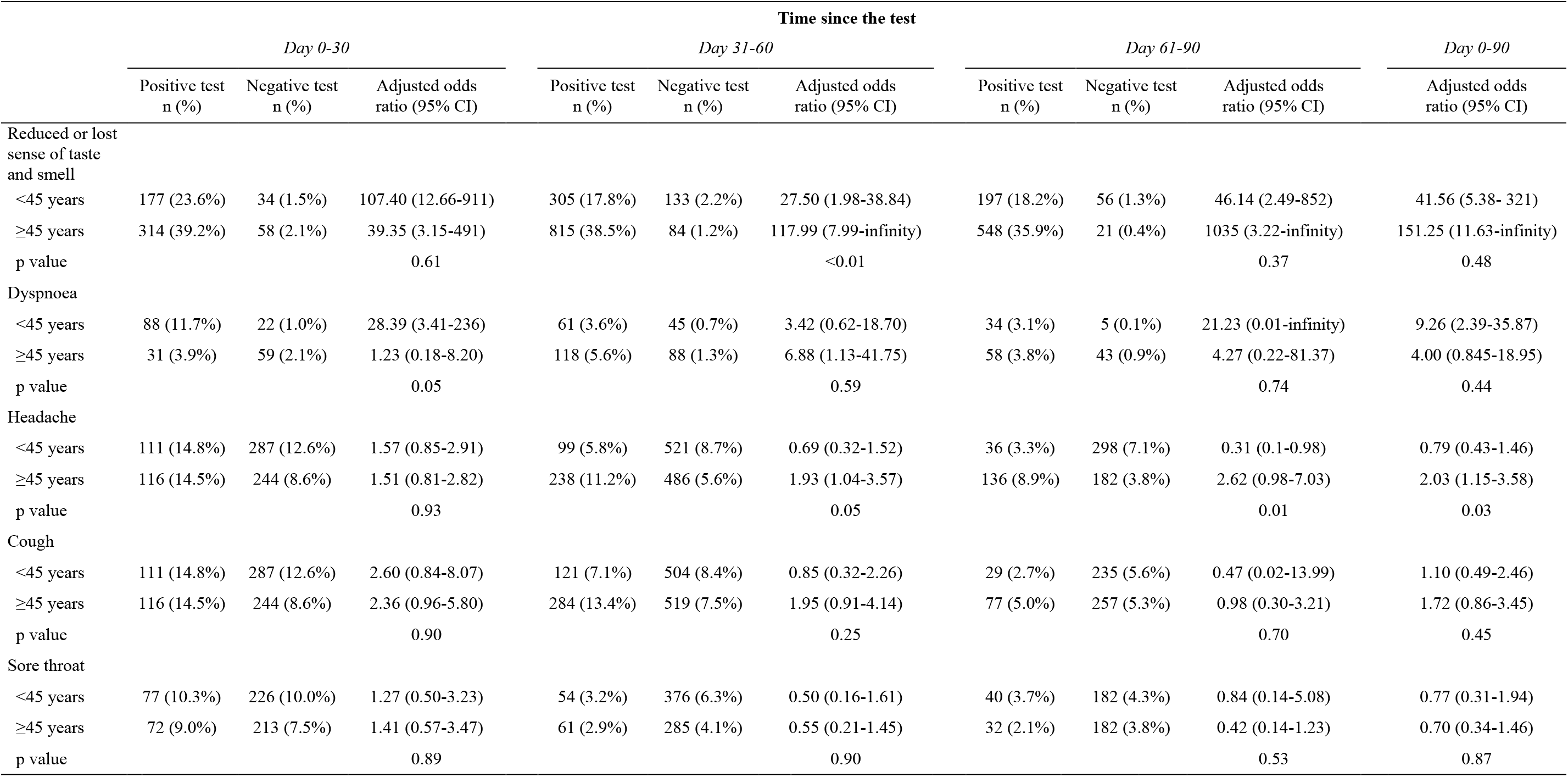
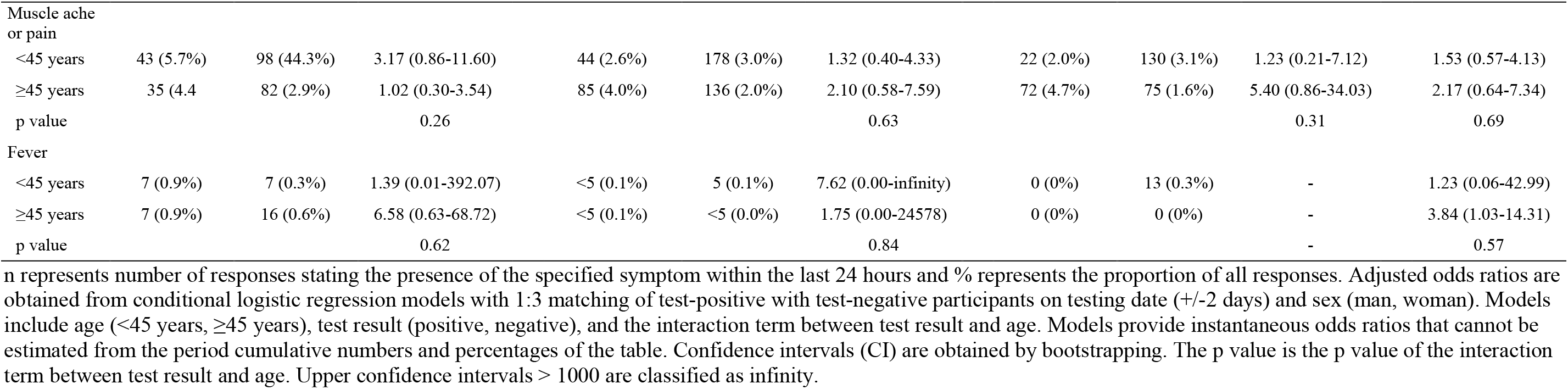
Odds ratios of seven symptoms by SARS-CoV-2 PCR test result, age, and time since the test.

|  | Time since the test |  |  |  |  |  |  |  |  |  |
| --- | --- | --- | --- | --- | --- | --- | --- | --- | --- | --- |
|  | Day 0-30 |  |  | Day 31-60 |  |  | Day 61-90 |  |  | Day 0-90 |
|  | Positive test<br>n (%) | Negative test<br>n (%) | Adjusted odds<br>ratio (95% CI) | Positive test<br>n (%) | Negative test<br>n (%) | Adjusted odds<br>ratio (95% CI) | Positive test<br>n (%) | Negative test<br>n (%) | Adjusted odds<br>ratio (95% CI) | Adjusted odds<br>ratio (95% CI) |
| Reduced or lost<br>sense of taste<br>and smell |  |  |  |  |  |  |  |  |  |  |
| <45 years | 177 (23.6%) | 34 (1.5%) | 107.40 (12.66-911) | 305 (17.8%) | 133 (2.2%) | 27.50 (1.98-38.84) | 197 (18.2%) | 56 (1.3%) | 46.14 (2.49-852) | 41.56 (5.38- 321) |
| ≥45 years | 314 (39.2%) | 58 (2.1%) | 39.35 (3.15-491) | 815 (38.5%) | 84 (1.2%) | 117.99 (7.99-infinity) | 548 (35.9%) | 21 (0.4%) | 1035 (3.22-infinity) | 151.25 (11.63-infinity) |
| p value |  |  | 0.61 |  |  | <0.01 |  |  | 0.37 | 0.48 |
| Dyspnoea |  |  |  |  |  |  |  |  |  |  |
| <45 years | 88 (11.7%) | 22 (1.0%) | 28.39 (3.41-236) | 61 (3.6%) | 45 (0.7%) | 3.42 (0.62-18.70) | 34 (3.1%) | 5 (0.1%) | 21.23 (0.01-infinity) | 9.26 (2.39-35.87) |
| ≥45 years | 31 (3.9%) | 59 (2.1%) | 1.23 (0.18-8.20) | 118 (5.6%) | 88 (1.3%) | 6.88 (1.13-41.75) | 58 (3.8%) | 43 (0.9%) | 4.27 (0.22-81.37) | 4.00 (0.845-18.95) |
| p value |  |  | 0.05 |  |  | 0.59 |  |  | 0.74 | 0.44 |
| Headache |  |  |  |  |  |  |  |  |  |  |
| <45 years | 111 (14.8%) | 287 (12.6%) | 1.57 (0.85-2.91) | 99 (5.8%) | 521 (8.7%) | 0.69 (0.32-1.52) | 36 (3.3%) | 298 (7.1%) | 0.31 (0.1-0.98) | 0.79 (0.43-1.46) |
| ≥45 years | 116 (14.5%) | 244 (8.6%) | 1.51 (0.81-2.82) | 238 (11.2%) | 486 (5.6%) | 1.93 (1.04-3.57) | 136 (8.9%) | 182 (3.8%) | 2.62 (0.98-7.03) | 2.03 (1.15-3.58) |
| p value |  |  | 0.93 |  |  | 0.05 |  |  | 0.01 | 0.03 |
| Cough |  |  |  |  |  |  |  |  |  |  |
| <45 years | 111 (14.8%) | 287 (12.6%) | 2.60 (0.84-8.07) | 121 (7.1%) | 504 (8.4%) | 0.85 (0.32-2.26) | 29 (2.7%) | 235 (5.6%) | 0.47 (0.02-13.99) | 1.10 (0.49-2.46) |
| ≥45 years | 116 (14.5%) | 244 (8.6%) | 2.36 (0.96-5.80) | 284 (13.4%) | 519 (7.5%) | 1.95 (0.91-4.14) | 77 (5.0%) | 257 (5.3%) | 0.98 (0.30-3.21) | 1.72 (0.86-3.45) |
| p value |  |  | 0.90 |  |  | 0.25 |  |  | 0.70 | 0.45 |
| Sore throat |  |  |  |  |  |  |  |  |  |  |
| <45 years | 77 (10.3%) | 226 (10.0%) | 1.27 (0.50-3.23) | 54 (3.2%) | 376 (6.3%) | 0.50 (0.16-1.61) | 40 (3.7%) | 182 (4.3%) | 0.84 (0.14-5.08) | 0.77 (0.31-1.94) |
| ≥45 years | 72 (9.0%) | 213 (7.5%) | 1.41 (0.57-3.47) | 61 (2.9%) | 285 (4.1%) | 0.55 (0.21-1.45) | 32 (2.1%) | 182 (3.8%) | 0.42 (0.14-1.23) | 0.70 (0.34-1.46) |
| p value |  |  | 0.89 |  |  | 0.90 |  |  | 0.53 | 0.87 |
|  | Day 0-30 |  |  | Day 31-60 |  |  | Day 61-90 |  |  | Day 0-90 |
|  | Positive test<br>n (%) | Negative test<br>n (%) | Adjusted odds<br>ratio (95% CI) | Positive test<br>n (%) | Negative test<br>n (%) | Adjusted odds<br>ratio (95% CI) | Positive test<br>n (%) | Negative test<br>n (%) | Adjusted odds<br>ratio (95% CI) | Adjusted odds<br>ratio (95% CI) |
| Muscle ache<br>or pain |  |  |  |  |  |  |  |  |  |  |
| <45 years | 43 (5.7%) | 98 (44.3%) | 3.17 (0.86-11.60) | 44 (2.6%) | 178 (3.0%) | 1.32 (0.40-4.33) | 22 (2.0%) | 130 (3.1%) | 1.23 (0.21-7.12) | 1.53 (0.57-4.13) |
| ≥45 years | 35 (4.4) | 82 (2.9%) | 1.02 (0.30-3.54) | 85 (4.0%) | 136 (2.0%) | 2.10 (0.58-7.59) | 72 (4.7%) | 75 (1.6%) | 5.40 (0.86-34.03) | 2.17 (0.64-7.34) |
| p value |  |  | 0.26 |  |  | 0.63 |  |  | 0.31 | 0.69 |
| Fever |  |  |  |  |  |  |  |  |  |  |
| <45 years | 7 (0.9%) | 7 (0.3%) | 1.39 (0.01-392.07) | <5 (0.1%) | 5 (0.1%) | 7.62 (0.00-infinity) | 0 (0%) | 13 (0.3%) | - | 1.23 (0.06-42.99) |
| ≥45 years | 7 (0.9%) | 16 (0.6%) | 6.58 (0.63-68.72) | <5 (0.1%) | <5 (0.0%) | 1.75 (0.00-24578) | 0 (0%) | 0 (0%) | - | 3.84 (1.03-14.31) |
| p value |  |  | 0.62 |  |  | 0.84 |  |  | - | 0.57 |

n represents number of responses stating the presence of the specified symptom within the last 24 hours and % represents the proportion of all responses. Adjusted odds ratios are obtained from conditional logistic regression models with 1:3 matching of test-positive with test-negative participants on testing date (+/-2 days) and sex (man, woman). Models include age (<45 years, ≥45 years), test result (positive, negative), and the interaction term between test result and age. Models provide instantaneous odds ratios that cannot be estimated from the period cumulative numbers and percentages of the table. Confidence intervals (CI) are obtained by bootstrapping. The p value is the p value of the interaction term between test result and age. Upper confidence intervals > 1000 are classified as infinity.

**Table S2:**
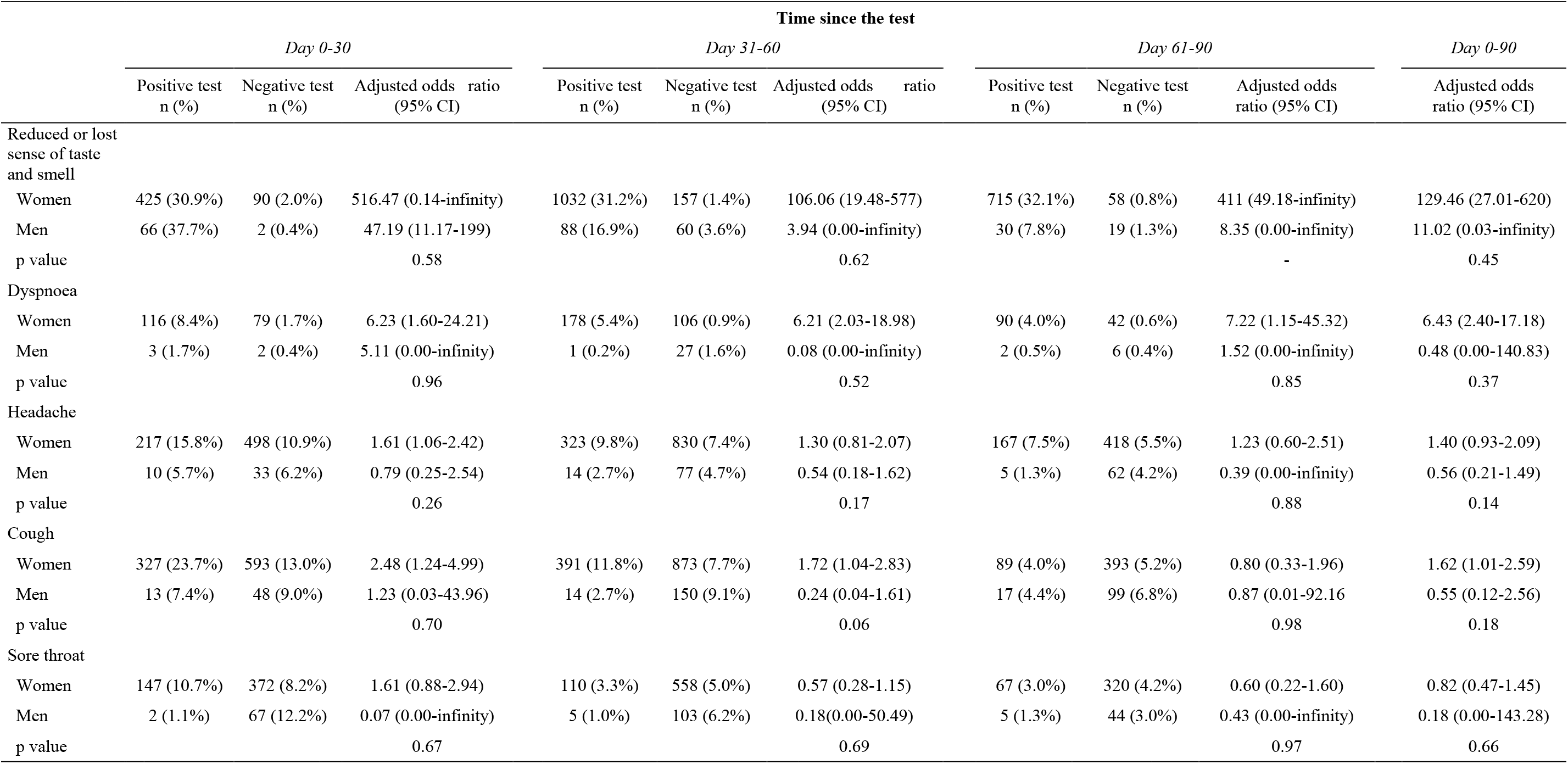
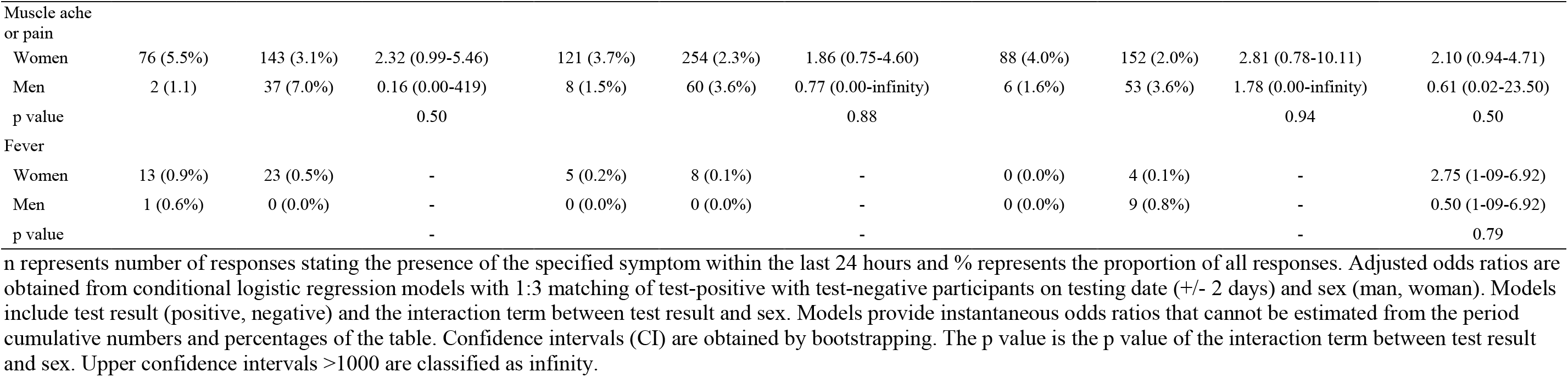
Odds ratios of seven symptoms by SARS-CoV-2 PCR test result, sex, and time since the test.

**Table S3:** Distribution of sex, age, and occupation from source population to final study population.

|  | Source population<sup>a</sup> | Participants reporting daily symptoms | Participants reporting daily symptoms and SARS-CoV-2 PCR tested | Final study population<sup>b</sup> |
| --- | --- | --- | --- | --- |
|  | n (%) | n (%) | n (%) | n (%) |
| Sex |  |  |  |  |
| Female | 25 524 (78.7%) | 10 232 (84.5%) | 3174 (87.5%) | 708 (84.3%) |
| Male | 6889 (21.3%) | 1883 (15.5%) | 453 (12.5%) | 132 (15.7%) |
| Age, years |  |  |  |  |
| <30 | 5833 (18.0%) | 1276 (10.5%) | 392 (10.8%) | 91 (10.8%) |
| 30-39 | 7281 (22.5%) | 2586 (21.3%) | 864 (23.8%) | 202 (24.1%) |
| 40-49 | 7822 (24.1%) | 3436 (28.4%) | 1079 (29.7%) | 285 (33.9%) |
| 50-59 | 7405 (22.8%) | 3373 (27.8%) | 909 (25.1%) | 195 (23.2%) |
| ≥60 | 4072 (12.6%) | 1444 (11.9%) | 383 (10.6%) | 67 (8.0%) |
| Occupation |  |  |  |  |
| Nursing staff | 11 556 (35.7%) | 4735 (39.1%) | 1614 (44.5%) | 430 (51.2%) |
| Medical doctors | 4615 (14.2%) | 1328 (11.0%) | 475 (13.1%) | 149 (17.7%) |
| Biomedical laboratory scientists | 1154 (3.6%) | 661 (5.5%) | 196 (5.4%) | 45 (5.4%) |
| Medical secretaries | 1904 (5.9%) | 1015 (8.4%) | 305 (8.4%) | 44 (5.2%) |
| Other | 13 184 (40.7%) | 4376 (36.1%) | 1037 (28.6%) | 172 (20.5%) |
| All | 32 413 (100%) | 12 115 (100%) | 3624 (100%) | 840 (100%) |

<sup>a</sup> All current employees in Central Denmark Region, March 31, 2020.
<sup>b</sup> Final study population including 210 SARS-CoV-2 test-positive participants and 630 3:1 sex and testing date (+/-2 days) matched random sample of test-negative participants.

## The COBRA-questionnaire

### COVID-19 among health care workers in Denmark (in Danish: COVID-19 blandt Regionsansatte)

The questionnaire includes a base line questionnaire filled in only once and focusing on the time since mid-February, 2020, where the first COVID-19 case was reported in Denmark, and a daily questionnaire focusing on the last 24 hours. Data collection started April 24, 2020 and ended June 30, 2020. Participants fill in their responses on their smartphone or computer. Every afternoon they receive a message with a link to the questionnaire available at REDCap electronic data capture software. The questionnaire data can be linked by the participants’ personal ID-numbers with SARS-CoV-2 PCR test results and other register data.

### Base line questions

1. **Have you had one or more of the following symptoms for 4 days or more since mid-February? (More tics if relevant)**
  a. Cough
  b. Sore throat
  c. Headache
  d. Muscle ache/pain
  e. Fever
  f. Dyspnea
  g. Reduced or loss of sense of taste and smell
  h. None of these
2. **Have you contacted a doctor because of these symptoms?**
  a. No
  b. Yes
3. **Are there any other in your household that have experienced cough, sore throat, headache, muscle pain, fever, and dyspnea or reduced or loss of sense of taste and smell for 4 days or more since mid-February?**
  a. No
  b. Yes
4. **Are you smoking?**
  a. Yes
  b. No, former smoker
  c. Never smoking

### Daily questions

1. **Have you had one or more of the following symptoms within the last 24 hours? (More tics if relevant)**
  a. Cough
  b. Sore throat
  c. Headache
  d. Muscle ache/pain
  e. Fever
  f. Dyspnea
  g. Reduced or loss of sense of taste and smell
  h. None of these
  *If responses a-g in question 1:* 1a) Have you contacted a doctor within the last 24 hours because of these symptoms?
  a. No
  b. Yes
2. **Are there any other in your household that have experienced cough, sore throat, headache, muscle pain, fever, and dyspnea or reduced or loss of senses of taste and smell within the last 24 hours?**
  a. No
  b. Yes
  c. Don’t know
3. **Have you undertaken any of the following tasks within the last 24 hours? (More tics if relevant)**
  a. Day off /sick/at home
  b. Consultations with patients within a 2 meters distance
  c. Physical contact with patients (e.g. treatment, examination, personal care, patient transfer)
  d. Surgical procedures or birth giving
  e. Procedures in airways (e.g. CPAP, PEP, intubation or resuscitation)
  f. Patient transport
  g. Other tasks within a 2 meters distance
  h. Preparation of hospital ward or cleaning
  i. I have been at work, but did not engage in any of the mentioned work tasks
  *If positive responses b-h in question 3:*
4. **Have you during work within the last 24 hours been in contact with subjects with suspected COVID-19 or tested positive for COVID-19?**
  a. No
  b. Yes, subjects tested positive for COVID-19
  c. Yes, subjects with suspected COVID-19
  d. Don’t know
  *If positive responses b-h in question 3:*
5. **What type of personal protective equipment have you used for the last 24 hours? (More tics if relevant)**
  a. I have not used personal protective equipment
  b. Gloves
  c. Gown with long sleeves
  d. Plastic apron
  e. High isolation gown
  f. Surgical mask-type IIR
  g. Respirator-type FFP2
  h. Respirator-type FFP3
  i. Respirator-unknown type
  j. Face shield
  k. Surgical mask with shield
  l. Protective glasses
  m. Other protective equipment
  *If response m in question 5:* Please specify what kind of protective equipment:________________ *If responses b-m in question 5:* 5a) Have there been any accidents with this protective equipment within the last 24 hours? (e.g. broken glove, dropped face shield or respirator)
  a. Yes
  b. No
  c. Don’t know
  *If response a in question 5a:* 5b) Which personal protective equipment were involved in the accident? *[Only those PPEs ticked in question 5 are presented for the participant]*
  a. Gloves
  b. Gown with long sleeves
  c. Plastic apron
  d. High isolation gown
  e. Surgical mask-type IIR
  f. Respirator-type FFP2
  g. Respirator-type FFP3
  h. Respirator-unknown type
  i. Face shield
  j. Surgical mask with shield
  k. Protective glasses
  l. Other protective equipment
  *If response l in question 5b:* Please specify what kind of protective equipment:____________ *If response a in question 5a:* 5c) During which task did the accident with your protective equipment happen? (More tics if relevant) *[Only the below mentioned tasks ticked in question 3 are presented for the participant]*
  a. Consultations with patients within a 2 meters distance
  b. Physical contact with patients (e.g. treatment, examination, personal care, patient transfer)
  c. Surgical procedures or birth giving
  d. Procedures in airways (e.g. CPAP, PEP, intubation or resuscitation)
  e. Patient transport
  f. Other tasks within a 2 meters distance
  g. Preparation of hospital ward or cleaning
  *If responses f in question 5c:* Describe the task during which you experienced the accident:___________ *If response a in question 5a:* Describe how the accident happened:___________ *If responses b or c in question 4:* 5d) Did the accident with personal protective equipment involve subjects under suspicion of or tested positive for COVID-19?
  a. No
  b. Yes, subjects tested positive for COVID-19
  c. Yes, subjects suspected of COVID-19
  d. Don’t know
6. **Has there within the last 24 hours been situations where you did not use the recommended personal protective equipment?**
  a. No
  b. Yes
  c. Don’t know
  *If response b in question 6:* 6a) Which recommended personal protective equipment did you not use? (More tics if relevant)
  a. Gloves
  b. Gown with long sleeves
  c. Plastic apron
  d. High isolation gown
  e. Surgical mask-type IIR
  f. Respirator-type FFP2
  g. Respirator-type FFP3
  h. Respirator-unknown type
  i. Face shield
  j. Surgical mask with shield
  k. Protective glasses
  l. Other protective equipment
  *If response l in question 6a:* Please specify what kind of protective equipment:____________ *If response b in question 6:* 6b) During which tasks did you not use the recommended personal protective equipment? (More ticks if relevant) *[Only the below mentioned tasks ticked in question 6a are presented for the participant]*
  a. Consultations with patients within a 2 meter distance
  b. Physical contact with patients (e.g. treatment, examination, personal care, patient transfer)
  c. Surgical procedures or birth giving
  d. Procedures in airways (e.g. CPAP, PEP, intubation or resuscitation)
  e. Patient transport
  f. Other tasks within a 2 meter distance
  g. Preparation of hospital ward or cleaning
  *If response f in question 6b:* *Please specify the other task:*_______________ *If response b or c in question 4 and response b in question 6:* 6c) Did you work with subjects under suspicion of or tested positive for COVID-19?
  a. No
  b. Yes, subjects tested positive for COVID-19
  c. Yes, subjects with suspected COVID-19
  d. Don’t know
  *If response b in question 6:* 6d) What was the reason for not using protective equipment? (More tics if relevant)
  a. Forgot it
  b. The personal protective equipment was not available
  c. Did not have time for it
  d. To spare equipment
  e. Unaware that I should use equipment
  f. Other reason
  *If response f in question 6d:* Please specify the reason for not using the recommended personal protective equipment: __________
7. **We appreciate your participation in the study. You will receive this questionnaire daily until June 30. Do you want to stop your participation now, please tic here?**
  a. I wish to stop now
  *If response a in question 7:* If you are sure you want to stop now, please confirm here:
  a. Yes, I wish to stop now
  b. No, I do not wish to stop now

